# AI vs Human Performance in Conversational Hospital-Based Neurological Diagnosis

**DOI:** 10.1101/2025.08.13.25333529

**Authors:** Moran Sorka, Alon Gorenshtein, Hillel Abramovitch, Pannathat Soontrapa, Shahar Shelly, Dvir Aran

**Author notes:** Corresponding authors: Dvir Aran, 21 Sderot Rose, Technion-Israel Institute of Technology, Haifa, Israel, 3095230, Shahar Shelly, Department of Neurology, Rambam Medical Center, Haifa, Israel.

## Abstract

**Background:** Most evaluations of artificial intelligence (AI) in medicine rely on static, multiple-choice benchmarks that fail to capture the dynamic, sequential nature of clinical diagnosis. While conversational AI has shown promise in telemedicine, these systems rarely test the iterative decision-making process in which clinicians gather information, order tests, and refine diagnoses.

**Methods:** We developed DiagnosticXchange, a web-based platform simulating realistic clinical interactions between providers and specialist consultants. A ‘nurse’ agent responds to requests from human physicians or AI systems acting as diagnosticians. Sixteen neurological diagnostic challenges of varying complexity were drawn from diverse educational and peer-reviewed sources. We evaluated 14 neurologists at different training stages and multiple state-of-the-art large language models (LLMs) using efficiency metrics, including: diagnostic accuracy, procedural cost efficiency (based on CPT codes and hospital pricing), and time to diagnosis (using actual procedure durations). We also developed Gregory, a specialized multi-agent system that systematically generates differential diagnoses, challenges initial hypotheses, and strategically selects high-yield diagnostic tests.

**Results:** Human neurologists achieved 81% diagnostic accuracy (79% residents, 88% specialists) across 97 sessions; base LLMs ranged from 81-94%. Gregory achieved perfect diagnostic accuracy with markedly lower diagnostic costs (average $1,423; 95% CI: $450-$2,860) compared with human neurologists (average $3,041; 95% CI: $2,464-$3,677; p=0.008) and base LLMs (average $2,759; 95% CI: $2,137-$3,476; p=0.002). Time to diagnosis was also shorter with Gregory (23 days; 95% CI: 6-48) versus human neurologists (43 days; 95% CI: 31-58; p=0.002) and base models (41 days; 95% CI: 31-51; p=0.07). The platform revealed distinct diagnostic patterns: human users and some base LLMs frequently ordered broad and expensive testing, while Gregory employed targeted strategies that avoided unnecessary procedures without sacrificing thoroughness.

**Conclusions:** A well-designed multi-agent AI system outperformed both human physicians and base LLMs in diagnostic accuracy, while reducing costs and time. DiagnosticXchange enables systematic evaluation of diagnostic efficiency and reasoning in realistic, interactive scenarios, offering a clinically relevant alternative to static benchmarks and a pathway toward more effective AI-assisted diagnosis.

## Introduction

Recent advances in large language models (LLMs) have shown remarkable performance on medical examinations,^1–5^ yet these assessments do not reflect how physicians actually practice medicine through sequential information gathering, hypothesis testing, and diagnostic refinement based on emerging evidence.^6^ Real-world diagnosis involves complex considerations, including diagnostic test costs that can range from hundreds to thousands of dollars, time delays that may span days to weeks, and resource constraints that force difficult trade-offs between thoroughness and efficiency. Physicians diagnosing a complex neurological condition may order dozens of tests, revise hypotheses multiple times, and spend weeks gathering information before reaching a conclusion. However, most AI evaluation in medicine reduces this intricate process to answering multiple-choice questions in isolation. This approach risks overstating model competence and obscures potential weaknesses including premature diagnostic closure, indiscriminate test ordering, and anchoring on early hypotheses.^7^ For instance, current benchmarks cannot capture whether an AI system would order unnecessary expensive imaging, fail to follow up on abnormal lab results, or anchor prematurely on an initial hypothesis. While useful for assessing medical knowledge, these static benchmarks fail to capture the dynamic and iterative nature of clinical diagnosis.

Recognizing these limitations, recent work has begun to address this gap by developing more dynamic evaluation approaches. However, early attempts have revealed significant challenges. Hager et al. demonstrated that LLMs performed significantly worse than physicians in iterative diagnostic tasks, failing to follow guidelines and showing sensitivity to information order and quantity.^8^ While these findings highlighted important limitations, the study used older AI models and relatively simple evaluation frameworks that may not reflect the full potential of current AI systems or the complexity of real clinical workflows. Google’s AMIE system represented a significant advance, demonstrating superior performance to primary care physicians in text-based consultations.^7^ However, this work was limited to telemedicine interactions, which represent only a narrow subset of medical practice. Most clinical encounters involve complex workflows where healthcare providers must coordinate with specialists, order diagnostic tests, interpret results, and make iterative decisions based on evolving clinical pictures. These realistic clinical workflows, where diagnosticians must systematically gather information and strategically select tests to reach conclusions, have not been adequately evaluated in current AI benchmarks.

What would an ideal AI diagnostic evaluation look like? It would need to capture the iterative trial-and-error process where initial treatments may fail, requiring diagnostic refinement and alternative approaches.^9,10^ It would assess not just accuracy but also cost-effectiveness, time efficiency, and resource optimization. Most critically, it would evaluate AI systems’ ability to reason transparently, quantify uncertainty, and avoid cognitive biases that compromise clinical decision-making.

Current autonomous AI agents often operate as black boxes without transparent reasoning and employ reactive rather than strategic planning approaches.^8,11^ Recent advances in multi-agent AI systems offer promising approaches to address these challenges by decomposing complex diagnostic reasoning into specialized cognitive functions.^10,12^ These frameworks can mirror expert physician reasoning through systematic differential diagnosis generation, hypothesis testing, and strategic test selection,^13,14^ while implementing structured processes that include evidence integration, bias detection, and cost-conscious decision-making.^15^ However, their effectiveness in realistic clinical workflows requiring iterative information gathering and resource optimization has been less extensively studied.^16^

Neurology presents an ideal testing ground for advanced AI evaluation due to its inherent complexity and the critical shortage of specialists worldwide.^17,18^ With fewer than 18,000 practicing neurologists in the United States serving a population of over 340 million, many hospitals lack adequate neurological expertise, particularly in rural and underserved areas. Neurological diagnoses often require integration of detailed anatomical knowledge, temporal pattern recognition, and synthesis across multiple neural systems.^19^ The field’s diagnostic complexity ranges from pattern recognition in common conditions like migraine to systematic exclusion approaches for rare disorders like autoimmune encephalitis, requiring fundamentally different reasoning strategies that thoroughly test AI capabilities. These challenges make neurology an ideal domain for testing advanced AI diagnostic capabilities, particularly in specialist consultation scenarios where complex cases require systematic evaluation and strategic test selection. The combination of diagnostic complexity, specialist shortage, and economic pressures creates an urgent need to evaluate whether AI systems can not only match human diagnostic accuracy but do so while optimizing resource utilization and time efficiency in realistic clinical workflows.

## Methods

### Platform Development

We developed DiagnosticXchange (http://diagnostixange.aran-lab.com/), a web-based application designed to simulate realistic clinical workflows involving information gathering agents and diagnostic decision-makers. The platform architecture consists of two primary components: (1) an information gathering agent with access to complete case information from processed clinical vignettes that responds to specific requests for patient history, physical examination findings, and diagnostic test results, and (2) a diagnostic decision-maker role filled by either human neurologists or AI systems acting as consultants who must gather information systematically to reach a diagnosis and treatment plan.

The information-gathering agent was programmed to provide only the information explicitly requested, avoiding unsolicited details that might bias the diagnostic process. When a participant requested information absent from the source case, the agent either returned a “partial information available” message (26.0% of all messages) or generated a “clinically plausible” synthetic response consistent with the overall case context (25.2%). Such synthesis was necessary to preserve conversational flow and to simulate the reality that clinicians often must fill in missing details from indirect sources. All synthetic responses were explicitly flagged in the log to enable transparency and post hoc analysis.

Each diagnostic session was limited to a maximum of 5 diagnostic attempts and 50 total messages to maintain realistic session durations and prevent indefinite exploration. Sessions were classified as successful if participants reached the correct diagnosis within these limits, and as failed if they exceeded the attempt limits or were unable to reach a correct conclusion. The platform tracks all requests made, responses provided, and the final diagnostic and therapeutic recommendations.

#### Case Selection and Processing

We curated 16 neurological diagnostic clinical challenges representing varying levels of complexity from a range of sources, including NEJM, educational neurology journals, and other peer-reviewed case reports selected for clarity and completeness rather than rarity (**Supplementary Table 1**). Cases were classified by complexity (simple, moderate or complex) based on expert neurologist assessment, considering factors including: diagnostic rarity, number of required investigations, subspecialty expertise needed, and presence of atypical presentations or overlapping syndromes. Cases were selected to represent a wide range of neurological subspecialties including neuroimmunology (Anti-NMDAR encephalitis, multiple sclerosis), neuro-oncology (neurosarcoidosis), vascular neurology (cerebral infarction), movement disorders (Wilson’s disease), infectious neurology (cryptococcal meningitis), and critical care neurology (thrombotic thrombocytopenia).

The system employs a comprehensive automated medical case analysis pipeline to extract clinical information from PDF-formatted medical case reports, combining traditional document parsing with artificial intelligence to achieve comprehensive clinical data extraction and classification. Complete technical details of the text extraction, figure identification, and clinical data extraction processes are provided in the Supplementary Methods.

#### Application Architecture and User Interface

Upon authentication, users access a curated case library where each case presents a standardized interface displaying the case title and specialty classification. Each diagnostic session begins with presentation of only the structured History of Present Illness (HPI), providing users with patient demographics, chief complaint, symptom timeline, and relevant medical history initially presented in the case report. The application implements a progressive information disclosure system that controls access to additional clinical data throughout the diagnostic process.

Users must actively request specific information types (laboratory results, imaging studies, physical examination findings) through natural language interactions with the “Nurse” agent, simulating the clinical decision-making process of ordering appropriate diagnostic tests. The application implements a comprehensive message processing pipeline that handles all user communications through a structured five-step workflow: message reception and processing, automated message classification into medical information requests, diagnosis submissions, clarification responses, and general inquiries, category-specific processing with appropriate database queries and cost calculations, response generation following strict clinical fidelity protocols, and final response delivery with session updates (More information provided in **Supplementary Methods**).

### Gregory Multi-Agent System

We developed Gregory, a specialized multi-agent AI system designed to systematically generate differential diagnoses, challenge initial hypotheses, and strategically select high-yield diagnostic tests (**Supplementary Figure 1**). Gregory employs a three-agent architecture designed to mirror the specialized cognitive processes of expert neurologists, decomposing complex neurological reasoning into distinct cognitive functions while incorporating mathematical frameworks for optimal decision-making under uncertainty. Gregory agents utilize the OpenAI o1 model.

#### Main Orchestrator Agent

The orchestrator serves as the central coordination hub, managing the overall diagnostic session flow and facilitating communication between specialized agents. Key responsibilities include session initialization, response analysis, and decision execution. The orchestrator processes both clinical information and clarification requests, maintaining session state and tracking diagnostic attempts to prevent infinite loops.

#### Differential Diagnosis Manager Agent

This agent specializes in evidence accumulation and differential diagnosis maintenance, extracting structured clinical findings from natural language exchanges using LLM-based analysis. The agent classifies evidence as supporting or contradicting for each potential diagnosis, maintaining comprehensive differential diagnosis lists with confidence scores and evidence tracking. A key innovation is its batch processing approach to evidence integration, utilizing both LLM-based reasoning and retrieval-augmented generation (RAG) for robust initial differential diagnosis creation using embeddings from neurological textbooks.^20^

#### Strategic Diagnostic Framework Agent

This agent determines optimal diagnostic strategies through comprehensive multi-dimensional analysis, performing analysis of diagnostic tests by calculating information gain, strategic value, and cost-effectiveness for each potential investigation. The framework implements advanced cognitive bias mitigation strategies, systematically analyzing differential diagnosis lists for anchoring bias, confirmation bias, and premature closure, integrating these analyses into decision-making to ensure balanced diagnostic reasoning.

Gregory implements a structured five-phase diagnostic process that combines information theory with systematic clinical reasoning:

**Phase 1: Evidence Accumulation** - Clinical information extraction using LLM-based analysis of chat exchanges to identify structured clinical findings. The framework classifies findings as supporting or contradicting evidence for each potential diagnosis and assigns evidence weights based on clinical significance. Each piece of evidence contributes to the entropy reduction of the diagnostic space, where entropy 𝐻(𝑌) = −𝛴 𝑃(𝑦_!_) 𝑙𝑜𝑔 𝑃(𝑦_!_) quantifies uncertainty about the true diagnosis Y.
**Phase 2: Differential Diagnosis Management** - Initial differential diagnoses are generated through integration of LLM-based reasoning and RAG-based knowledge retrieval. The framework performs comprehensive differential updates using all accumulated evidence, applying Bayesian confidence adjustments where 𝑃(𝑑𝑖𝑎𝑔𝑛𝑜𝑠𝑖𝑠|𝑒𝑣𝑖𝑑𝑒𝑛𝑐𝑒) α 𝑃(𝑒𝑣𝑖𝑑𝑒𝑛𝑐𝑒|𝑑𝑖𝑎𝑔𝑛𝑜𝑠𝑖𝑠) × 𝑃(𝑑𝑖𝑎𝑔𝑛𝑜𝑠𝑖𝑠). Active differential tracking maintains lists of non-ruled-out diagnoses with probability distributions and supporting evidence.
**Phase 3: Strategic Test Analysis** - The system extracts all unique diagnostic tests from the differential diagnosis list and performs comprehensive analysis including cost analysis and evaluation of rule-out capabilities. Information gain 𝐼𝐺 = 𝐻(𝑌) − 𝐻(𝑌|𝑋) quantifies the expected reduction in diagnostic uncertainty from test X. Strategic value is calculated as: 𝑆𝑡𝑟𝑎𝑡𝑒𝑔𝑖𝑐 𝑉𝑎𝑙𝑢𝑒 = (𝐼𝐺 × 𝑇𝑟𝑒𝑎𝑡𝑎𝑏𝑖𝑙𝑖𝑡𝑦 × 𝑈𝑟𝑔𝑒𝑛𝑐𝑦) / (𝐶𝑜𝑠𝑡 × 𝑇𝑖𝑚𝑒_𝑃𝑒𝑛𝑎𝑙𝑡𝑦), where treatability bonus is 3.0× for conditions with available interventions, urgency bonus is 2.0× for time-sensitive conditions, and cost penalties range from 5 (low) to 15 (high) based on Current Procedural Terminology (CPT) code analysis.
**Phase 4: Cognitive Bias Analysis** - Systematic analysis of the differential diagnosis list identifies potential cognitive biases including anchoring bias, confirmation bias, and premature closure. Risk assessments quantify the potential impact of identified biases, with debiasing strategies integrated into decision-making processes.
**Phase 5: Strategic Decision Making** - Enhanced diagnosis submission criteria analysis determines whether sufficient evidence exists to submit a diagnosis or whether further investigation is warranted. The framework ranks tests by strategic score and selects optimal next test requests, considering invasiveness, treatability, cost-effectiveness, and avoidance of redundancy.

### Participants and AI System Evaluation

We recruited 14 neurologists at different training stages, including residents (PGY-1 through PGY-4) and attending physicians with varying years of neurological experience. Due to scheduling constraints and platform availability, not all participants completed all cases. We evaluated multiple state-of-the-art LLMs including GPT-4o, o1, Claude Sonnet-4, Gemini 2.5 Pro and Flash, comparing their performance against both human neurologists and Gregory across all 16 neurological cases. Participants received brief training on the platform interface before beginning their assigned cases.

### Outcome Measures

We evaluated performance across five metrics: diagnostic accuracy (primary outcome), cost efficiency, time to diagnosis, number of diagnostic attempts, and interaction length. The primary outcome was diagnostic accuracy, defined as the percentage of cases where the final diagnosis matched the ground truth diagnosis established in the original published case report. We emphasize that our analyses rely on relative differences between diagnosticians, not absolute values. Absolute costs will vary by payer and negotiated rates, but changes are expected to scale proportionally across providers, leaving relative performance rankings intact.

#### Cost Calculation

For each diagnostic request made during clinical sessions, we employed an LLM-based lookup system to translate diagnostic test requests into standardized CPT codes. These CPT codes were matched to corresponding cost data derived from negotiated rates for Anthem at Cleveland Clinic, sourced from hospital pricing transparency files under the Centers for Medicare & Medicaid Services (CMS) price transparency rule. Although cost estimates are not intended as exact representations of actual clinical expenses, they provide a standardized and consistent

#### Time Calculation

Time to diagnosis was calculated based on the cumulative duration required for all requested diagnostic procedures and tests. For each CPT code, time estimates reflecting real-world delays from test ordering to result availability were determined by an expert neurologist who was blinded to the diagnostic sessions and provided only the CPT codes and their descriptions. These estimates account for typical scheduling delays, procedure durations, result interpretation times, insurance approval processes, institutional protocols, and healthcare system administrative requirements encountered in clinical practice.

#### Additional Metrics

To analyze diagnostic test utilization patterns, CPT codes were grouped into clinically meaningful categories using the Clinical Classifications Software (CCS) framework. We also measured the total number of diagnostic attempts made before reaching final diagnosis and the total number of messages exchanged during each diagnostic session.

### Statistical Analysis

We compared performance metrics between human neurologists and AI systems using regression models that accounted for case-specific effects. Case-specific effects were included because participants completed different subsets of cases rather than all participants completing all cases, requiring statistical adjustment for the varying difficulty and characteristics of individual cases. For the primary analysis of diagnostic costs, we employed linear regression models with provider type and case ID as fixed effects to control for variations in case-specific difficulty. To address potential outliers and ensure robust results, we used robust regression methods via lmrob() from the robustbase package in R, which employs MM-estimators that are less sensitive to extreme values while providing standard p-values. For skewed continuous variables, we report the mean together with a 95% confidence interval derived from nonparametric bootstrap resampling (10,000 replicates). Statistical significance was set at p<0.05. All analyses were conducted in R (version 4.41).

### Ethics Statement

The Helsinki Committee (Ethics Committee) of Rambam Health Care Campus gave ethical approval for this work (Approval number: RMB-0026-24).

## Results

### Platform Development

We developed DiagnosticXchange to assess diagnostic performance through interactive clinical workflows (**Figure 1A**). The platform enables users to interact with complex neurological cases through a conversational interface where they request information, order diagnostic tests, and iteratively refine their diagnostic hypotheses. The system tracks every interaction, creating detailed diagnostic trajectories that reveal how different decision-makers approach complex clinical problems (**Figure 1B**). This comprehensive tracking capability allows evaluation beyond traditional accuracy metrics by measuring the associated costs and time required for each diagnostic pathway. The platform records every diagnostic request, calculates real-world costs using CPT codes and hospital pricing data, and estimates the time needed for each procedure and test result interpretation. This multidimensional assessment provides realistic evaluation of diagnostic performance that considers the practical constraints physicians face in clinical practice, where diagnostic decisions must balance accuracy against resource utilization and time efficiency.

**Figure 1.**
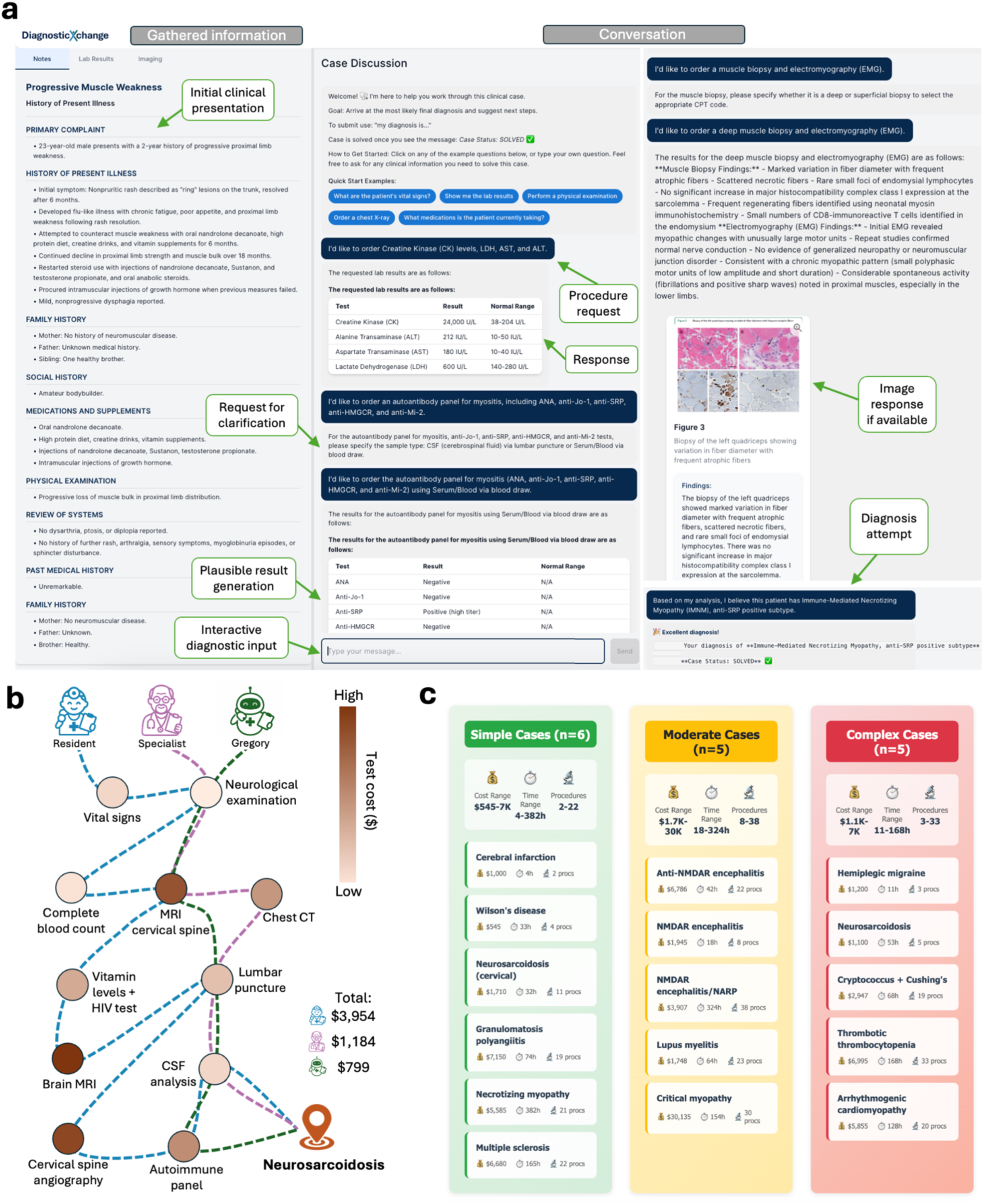
DiagnosticXchange Platform and Case Characteristics. **(A)** Screenshot of the DiagnosticXchange web-based platform interface. The left panel displays the initial case presentation with patient history, while the right panel shows the interactive conversation interface where diagnostic decision-makers request specific clinical information from a Nurse agent. Arrows indicate the request-response workflow that tracks all interactions and calculates costs using CPT codes. **(B)** Comparison of diagnostic trajectories between a resident physician (blue dashed line) and AI system (green dashed line) for a representative case involving a 48-year-old man with neurosarcoidosis. Each circle represents a diagnostic test or procedure, with size and color intensity proportional to cost (darker = more expensive). The AI system achieved the same correct diagnosis with lower total cost and reduced time through more targeted test selection. **(C)** Description of 16 neurological cases by complexity level used in the study. Simple cases (n=6), moderate cases (n=5, yellow) and complex cases (n=5). The final diagnosis is shown for each case spanning multiple neurological subspecialties. Cost and time presented are extracted from the procedures described in the original case.

### Cohort and Case Characteristics

We evaluated the platform using 16 neurological cases representing varying complexity levels across multiple subspecialties (**Figure 1C** and **Supplementary Table 1**). The cases span diverse neurological domains, requiring fundamentally different diagnostic approaches from pattern recognition in common conditions to systematic exclusion of rare disorders. Fourteen neurologists at different training stages completed a total of 97 diagnostic sessions across the 16 cases (**Supplementary Table 2**). Our neurologist cohort comprised residents (n=9) and attending physicians (n=5) with varying years of neurological experience. We compared human performance against multiple AI systems, including state-of-the-art LLMs and Gregory, our specialized multi-agent framework designed specifically for neurological reasoning.

### Diagnostic Performance

Across 193 total diagnostic sessions (human and AI), we observed clear differences in diagnostic accuracy between human neurologists and AI systems (**Figure 2A-D**). Human neurologists achieved varying success rates by experience level: residents 79% (56/71 sessions) and attending physicians 88% (23/26 sessions), with an overall human success rate of 83% (79/95 sessions). Base LLMs showed variable performance, with success rates ranging from 81% (GPT-4o) to 94% (o1 and Claude Sonnet-4). Notably, Gregory achieved perfect diagnostic accuracy, representing the only system to correctly diagnose all cases.

**Figure 2.**
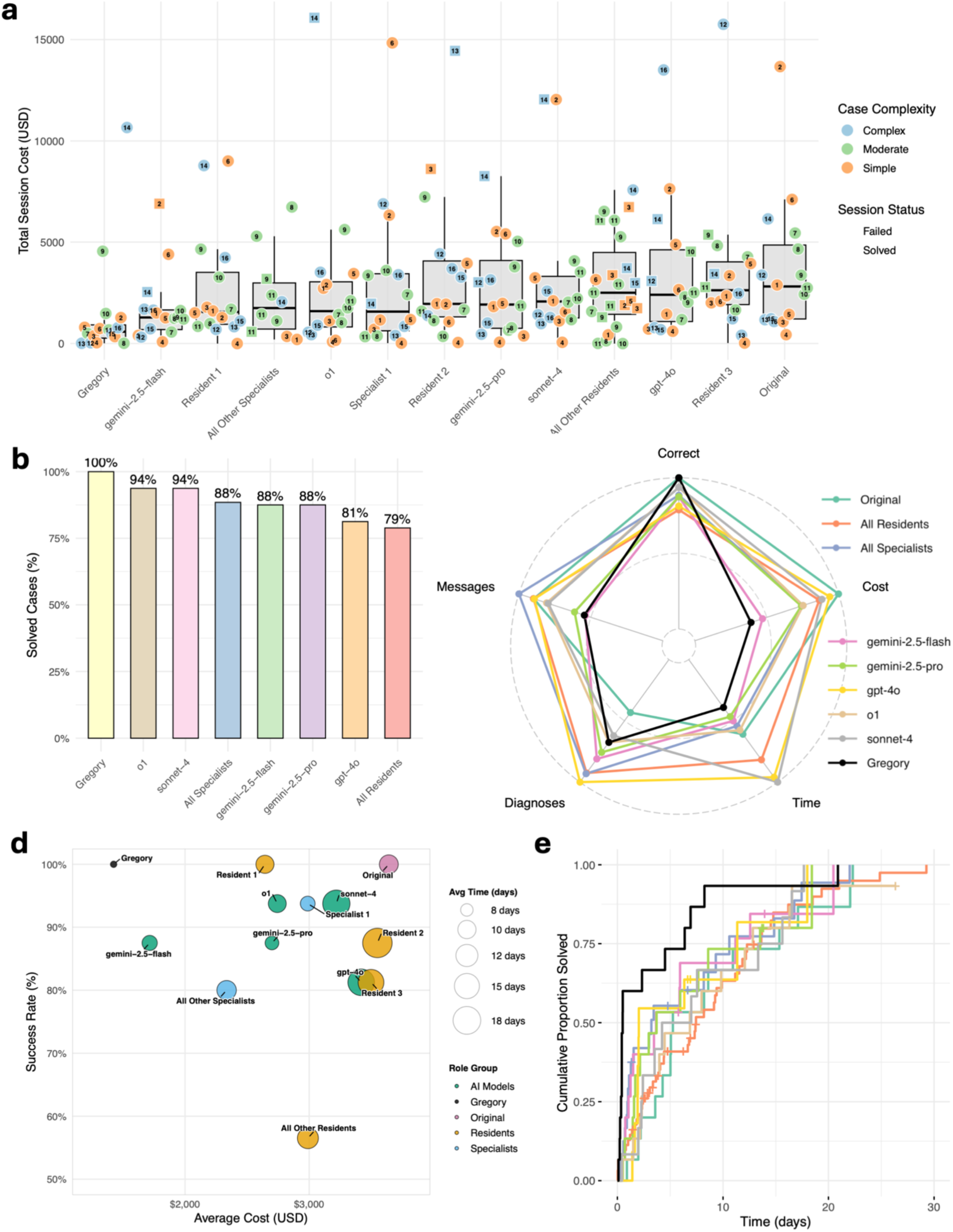
Diagnostic Performance and Efficiency Comparison Across AI Systems and Human Neurologists. **(A)** Individual session results showing total diagnostic costs (y-axis) for each provider type. Points represent individual diagnostic sessions colored by case complexity (green=complex, blue=moderate, orange=simple) and shaped by session outcome (circles=solved, triangles=failed). Box plots show median and interquartile ranges. **(B)** Diagnostic accuracy rates across different AI systems and provider types. **(C)** Radar plot comparing normalized performance across five metrics: diagnostic correctness, cost efficiency, time efficiency, number of diagnostic attempts, and message count. **(D)** Bubble plot of average diagnostic cost versus success rate, with bubble size representing average time to diagnosis. **(E)** Cumulative proportion of cases solved over time, showing Gregory’s faster diagnostic resolution compared to other providers.

Performance patterns revealed interesting case-specific variations that highlight the complementary strengths and weaknesses of human versus AI diagnostic approaches. In a complex case involving arrhythmogenic cardiomyopathy (Case #14), which presented with neurological symptoms despite having a primarily cardiac etiology, 4 out of 7 neurologists successfully identified the correct diagnosis, while all base LLMs failed to reach the correct diagnosis. Gregory successfully diagnosed this challenging case, demonstrating superior performance on diagnostically ambiguous presentations that span multiple medical domains. Conversely, in a case designated as simple involving cerebral peduncular infarction (Case #3), human neurologists showed similar performance with only 4 out of 7 reaching the correct diagnosis, but all base LLMs successfully identified this condition.

### Resource Efficiency Analysis

Cost efficiency analyses revealed dramatic differences between diagnostic approaches (**Figure 2A,C-D**). For reference, the original published cases had a total average cost of $3,642 per case (95% CI: $2,257–$5,414) based on all procedures performed and reported. Human neurologists demonstrated cost differences between training levels: residents averaged $3,152 per case (95% CI: $2,488–$3,901), while attending physicians averaged $2,759 per case (95% CI: $2,137–$3,476). Base LLMs showed variable costs, with Gemini-2.5-Flash being most cost-efficient among base models at average cost of $1,714 (95% CI: $1,020–$2,639), while GPT-4o was most expensive at $3,419 (95% CI: $2,013–$5,188) average cost. Gregory demonstrated superior cost-effectiveness with an average cost of $1,423 per case (95% CI: $450–$2,860), significantly lower than both human neurologists (p=0.008) and base LLMs (p=0.002). When comparing across human and AI models, Gregory achieved the lowest cost in 7 out of 16 cases (44%), ranked among the top three most cost-effective approaches in 11 out of 16 cases (69%), and demonstrated superior cost efficiency compared to other AI models in 12 out of 16 cases (75%).

Time efficiency analysis showed similar patterns (**Figure 2C-E**). The original published cases had an average diagnostic timeline of 35 days (95% CI: 22–51). Human providers required substantially longer: residents averaged 48 days (95% CI: 32–66) and attending physicians 32 days (95% CI: 15–52). Base AI models varied considerably in time efficiency, with Gemini-2.5-Pro achieving 27 days average time (95% CI: 15–41) but Claude Sonnet-4 requiring 58 days (95% CI: 31–89). Gregory achieved the fastest average diagnostic time at 23 days (95% CI: 6–48), representing a 36% improvement over the original case timelines and significantly faster than both human neurologists (p=0.002) and base LLM models (p=0.07).

Additional efficiency metrics further demonstrated Gregory’s systematic advantages. Gregory required fewer diagnostic attempts before reaching the correct conclusion: averaging 1.56 attempts per case compared to 1.79 for other AI models, 2.09 for residents, and 2.15 for attending physicians. Furthermore, Gregory demonstrated exceptional efficiency in diagnostic reasoning, achieving correct diagnoses with minimal information gathering. In 9 of 16 cases, Gregory required only 1-5 messages before reaching the correct diagnosis and completed the diagnostic process within half a day. In contrast, only 20% of all other sessions (human neurologists and base AI models combined) achieved diagnosis within 5 messages, and merely 10% completed diagnosis in less than half a day.

Overall, across our five key metrics (diagnostic accuracy, cost efficiency, time efficiency, number of diagnostic attempts, and interaction length), Gregory demonstrated a superior performance profile compared to both residents and attending physicians (**Figure 2C**). While some individual neurologists achieved high accuracy, they did so at substantially higher costs and longer times compared to Gregory. Base LLMs showed inconsistent performance with high variability in both accuracy and cost-effectiveness. Notably, when comparing Gregory directly to its underlying model (o1), Gregory outperformed o1 across all five metrics in every case, achieving 100% accuracy compared to o1’s 94% while reducing average costs by 48% and diagnostic time by 32%.

### Diagnostic Approach Patterns

Analysis of diagnostic trajectories revealed fundamental differences in reasoning strategies across experience levels and system types (**Figure 3A**). Training level differences emerged clearly between residents and specialists, with residents demonstrating more exploratory “shotgun” diagnostic approaches compared to specialists’ targeted strategies. Residents ordered more head CT scans (44.4% vs 3.6%) and non-standard laboratory tests (61.7% vs 46.4%), including extensive panels for dexamethasone suppression, various antibody screens, and infectious disease markers. In contrast, specialists showed more selective testing patterns while maintaining similar reliance on advanced imaging like MRI (65.4% vs 67.9%).

**Figure 3.**
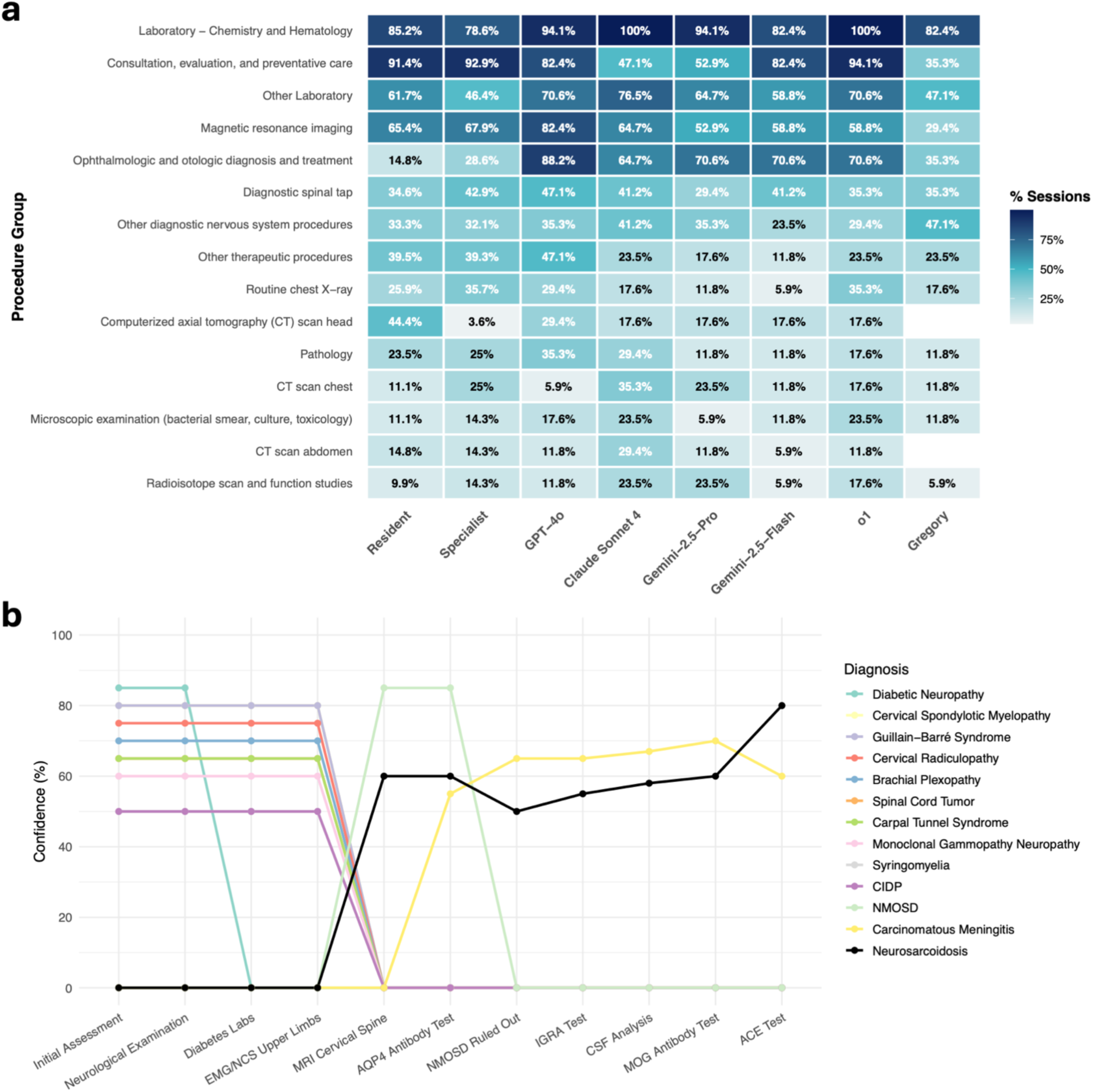
Diagnostic Reasoning Process and Test Utilization Patterns. **(A)** Diagnostic procedure utilization across provider types. Heatmap showing the percentage of diagnostic sessions that included each procedure group, comparing human neurologists with AI systems. Color intensity represents utilization frequency, with darker blue indicating higher usage rates. **(B)** Dynamic differential diagnosis evolution in Gregory’s neurosarcoidosis case. Confidence trajectories for all considered diagnoses across the diagnostic sequence, from initial clinical assessment through final ACE testing. Each colored line represents a different diagnostic hypothesis, with line thickness corresponding to peak confidence levels. The black line (neurosarcoidosis) shows Gregory’s systematic evidence accumulation leading to correct diagnosis, while other hypotheses are systematically evaluated and ruled out through strategic testing. Key decision points include the dramatic restructuring after cervical MRI (eliminating peripheral causes) and the confirmatory ACE elevation that established neurosarcoidosis as the final diagnosis at 80% confidence.

AI systems demonstrated remarkable heterogeneity in their diagnostic approaches, particularly in physical examination utilization. While some AI models closely mimicked human patterns with high reliance on physical examinations (o1 = 94.1% and GPT-4o = 82.4%) matching human levels (91-93%), others showed more selective approaches with Claude Sonnet-4 (47.1%) and Gemini-2.5-Pro (52.9%) using physical examinations in roughly half of sessions. Similarly, ophthalmologic and otologic evaluations, which primarily consist of sensorimotor examinations, were used minimally by humans (15-29%) but extensively by most AI systems (59-88%), suggesting different prioritization of neurological localization strategies. Gregory’s approach represented a distinctly precision-focused strategy that differed markedly from both human providers and base AI models.

While maintaining diagnostic accuracy, Gregory used fewer broad diagnostic modalities: MRI in only 29.4% of sessions compared to 53-82% for other systems, and physical examinations in 35.3% compared to 47-94% for others. Instead, Gregory demonstrated preferential use of targeted neurological procedures (47.1% vs 23-41% for other providers), including nerve conduction studies, evoked potentials, and specialized neurophysiological testing. This pattern suggests a strategic shift from broad information gathering toward precision diagnostic testing based on specific clinical hypotheses.

To illustrate Gregory’s systematic diagnostic approach, we examine a challenging neurosarcoidosis case that required extensive differential restructuring and dynamic hypothesis generation (**Figure 1B and Figure 3B).** A 48-year-old male presented with a six-week history of progressive “pins and needles” sensation beginning in the left hand and evolving to involve bilateral arms, upper chest, and posterior thighs, with symptoms exacerbated by neck hyperextension.

Gregory’s diagnostic process activated iteratively with each new piece of clinical information. Initial evaluation focused on peripheral causes, with diabetic neuropathy (85%), cervical spondylotic myelopathy (80%), and Guillain-Barré syndrome (80%) leading the differential, while central inflammatory conditions like neurosarcoidosis, NMOSD, and carcinomatous meningitis remained at 0% confidence. The critical paradigm shift occurred with cervical spine MRI revealing longitudinally extensive transverse myelopathy (LETM) from C4-C7 with leptomeningeal enhancement. Gregory’s Phase 2 triggered a complete restructuring: peripheral diagnoses were ruled out and NMOSD emerged as the leading hypothesis at 85% confidence, with neurosarcoidosis (60%) and carcinomatous meningitis also entering the differential. Gregory initially submitted NMOSD as its diagnosis, but when this proved incorrect, the system immediately restructured again. Carcinomatous meningitis rose to 65% confidence while neurosarcoidosis maintained significant probability. Subsequent testing with IGRA (reducing tuberculous meningitis from 30% to 15%) and CSF analysis revealing elevated protein (84 mg/dL) supported inflammatory processes. The addition of MOG antibody disease at 60% confidence demonstrated Gregory’s continued hypothesis generation, but when MOG antibodies returned negative, Phase 4 (Cognitive Bias Analysis) prevented anchoring on autoimmune demyelinating diseases.

The decisive iteration came with serum ACE testing, selected through Phase 3 (Key Tests Analysis) for its high information gain relative to cost. The elevated result (65 U/L) provided confirmatory evidence for neurosarcoidosis, which surged from moderate confidence to 80%, surpassing all other diagnoses. Phase 5 (Strategic Decision Making) determined that convergent evidence: progressive sensory symptoms, LETM pattern, leptomeningeal enhancement, elevated ACE, negative infectious and autoimmune workup, met criteria for diagnostic submission with final confidence of 85%.

Compared to this systematic trajectory, other providers displayed more variable and often less efficient approaches. Several neurologists began with appropriate initial testing but coupled it with broad simultaneous imaging and laboratory orders, raising cost without accelerating diagnostic narrowing; others anchored prematurely on demyelinating disease despite atypical features and either omitted or delayed systemic evaluation. Base AI models varied from prolonged initial laboratory workups to premature closure on inflammatory myelopathy without confirming systemic involvement, leading to missed or incomplete diagnoses.

## Discussion

This study introduces DiagnosticXchange, a novel evaluation framework that captures the iterative, resource-constrained nature of real clinical decision-making, addressing current limitations of existing AI evaluation approaches. Unlike static multiple-choice benchmarks that reduce diagnosis to pattern recognition, our platform simulates realistic clinical workflows where diagnostic decision-makers must systematically gather information, order tests, and refine hypotheses based on emerging evidence. The platform revealed distinct diagnostic patterns across experience levels and AI architectures, demonstrating its utility for both AI system evaluation and development. Our findings show that multi-agent AI systems can systematically exceed human diagnostic performance while reducing costs by 53% and diagnostic time by 47%, with Gregory achieving perfect diagnostic accuracy across 16 complex neurological cases while human neurologists averaged 83% accuracy at substantially higher resource costs.

Our approach takes current AI diagnostic evaluation one step forward. Microsoft’s SDBench used exclusively rare NEJM cases that don’t reflect real-world case distributions and allowed only single diagnostic attempts, unlike the iterative refinement typical of clinical practice.^21^ Google’s AMIE system, while demonstrating superior performance in telemedicine consultations,^7^ was limited to primary care interactions that lack the complex diagnostic workups and resource optimization challenges central to hospital-based specialist care. Previous studies raised concerns about AI readiness for clinical deployment. Hager et al. showed LLMs performing significantly worse than physicians in iterative tasks,^8^ but used older models with simple evaluation frameworks. DiagnosticXchange addresses these limitations by providing a comprehensive platform that evaluates not just diagnostic accuracy but also cost-effectiveness and time efficiency. Our work further demonstrates that well-designed multi-agent systems can overcome these limitations through systematic evidence integration, strategic test selection, and cognitive bias mitigation, capabilities absent in base LLMs.

Gregory’s superiority stems from its systematic decomposition of diagnostic reasoning into specialized cognitive functions. Unlike base LLMs that operate reactively, Gregory implements explicit information-theoretic optimization where each diagnostic action’s value is mathematically quantified. The multi-agent architecture prevents common diagnostic errors: anchoring bias through systematic differential restructuring, premature closure through explicit confidence thresholds, and resource waste through cost-benefit analysis integrated into every decision. The neurosarcoidosis case exemplifies this systematic approach. While human providers often anchored on initial hypotheses or ordered broad testing panels, Gregory’s evidence-driven differential restructuring and strategic test selection achieved the correct diagnosis efficiently. Importantly, this process is highly reproducible. Repeated runs on the same case produce nearly identical reasoning sequences, test choices, and final diagnoses. This stability contrasts with the variability observed in base LLMs and is critical for clinical adoption, where reproducibility and auditability are essential for trust.

It is important to note that Gregory is not a machine learning model trained on diagnostic cases, but rather a systematic multi-agent framework that orchestrates existing LLMs (OpenAI-o1 in this case) to optimize diagnostic reasoning. The consistent superiority over the base o1 model across all cases demonstrates that the value lies in the systematic approach to evidence gathering, differential diagnosis management, and strategic test selection, rather than memorization of specific cases.

Successful deployment requires integration with existing workflows and clear human-AI collaboration protocols. Gregory demonstrates readiness for specialist consultation scenarios where systematic diagnostic workups are standard practice. However, implementation must address current limitations: inability to perform physical examinations, need for physician oversight in complex cases, and integration with electronic health records. The economic implications are substantial. Gregory’s cost reductions could generate significant healthcare savings while addressing neurologist shortages through expert-level diagnostic capability in underserved areas. The reduction in diagnostic time represents faster treatment initiation and reduced patient anxiety in the diagnostic odyssey, clinically meaningful improvements beyond cost considerations.

## Data Availability

All data produced in the present study are available upon reasonable request to the authors.

## We acknowledge several limitations

First, the case set comprised only 16 neurological cases. This small number reflects the need to recruit human neurologists for head-to-head comparison, which constrained scalability. While this approach enabled a fair, directly comparable evaluation, it also limits statistical power and the breadth of clinical scenarios covered. Second, our focus on neurology was chosen for its diagnostic complexity and suitability as a “stress test” for reasoning systems, but this may limit generalizability to other specialties. Third, although our platform more closely mirrors clinical workflows than traditional static benchmarks, it remains a simulation. Real-world consult environments introduce factors not modeled here, including time pressure from concurrent responsibilities, incomplete or contradictory chart data, variable patient cooperation, and the need for physical examination. Fourth, a substantial proportion of queries involved partial or synthetic responses when requested details were not available in the original case. While these simulated missing-data events are a realistic feature of clinical encounters, they may have influenced diagnostic reasoning and should be considered when interpreting comparative performance. Finally, our cost and time estimates are based on a single payer–provider rate schedule and neurologist-derived timelines. These provide standardized comparisons but may differ from the conditions in other institutions.

Future work should prioritize prospective validation in real clinical environments, expansion to other medical domains, and larger case libraries that include common consult scenarios. The platform’s educational potential, providing standardized diagnostic challenges with detailed feedback, represents a promising application for training physicians in systematic reasoning and bias recognition. Integration with physical examination inputs and real-time patient interactions will be essential for broader applicability. Long-term studies examining clinical outcomes, physician acceptance, and healthcare economics will determine the practical impact of AI-assisted diagnosis. Research into optimal human– AI collaboration frameworks, in which AI augments rather than replaces physician decision-making, will be an important step toward safe and effective deployment.

In summary, a well-designed multi-agent AI system outperformed both human physicians and base LLMs in diagnostic accuracy, while reducing costs and time. Our web-based platform enables evaluation of AI systems in realistic clinical workflows that capture the complexity of actual medical practice. The systematic approach to diagnostic reasoning, with explicit bias mitigation and cost optimization, offers a path toward more effective and equitable healthcare delivery, though careful clinical validation and human-AI collaboration frameworks remain essential for successful implementation.

## Author Contributions

D.A., S.S., and M.S. conceived the study design and methodology. M.S. developed the DiagnosticXchange platform, implemented the Gregory multi-agent system, and wrote all code for data processing and analysis. D.A. performed the statistical analyses, created all figures and visualizations, and led the manuscript preparation. S.S. provided clinical expertise, reviewed case selections for neurological accuracy, and contributed clinical interpretation of results. A.G. supported clinical case curation and validation. D.A., S.S., and M.S. jointly wrote and revised the manuscript.

## Data Availability

All code, processed data, and materials used in this study are openly available at https://github.com/moransorka1/diagnosticXchange. The repository includes the DiagnosticXchange platform code, the Gregory multi-agent system, structured case data, and analysis scripts. Raw clinical cases used for simulation were derived from publicly available case reports.

## Supplementary Figures

**Supplementary Figure 1.**
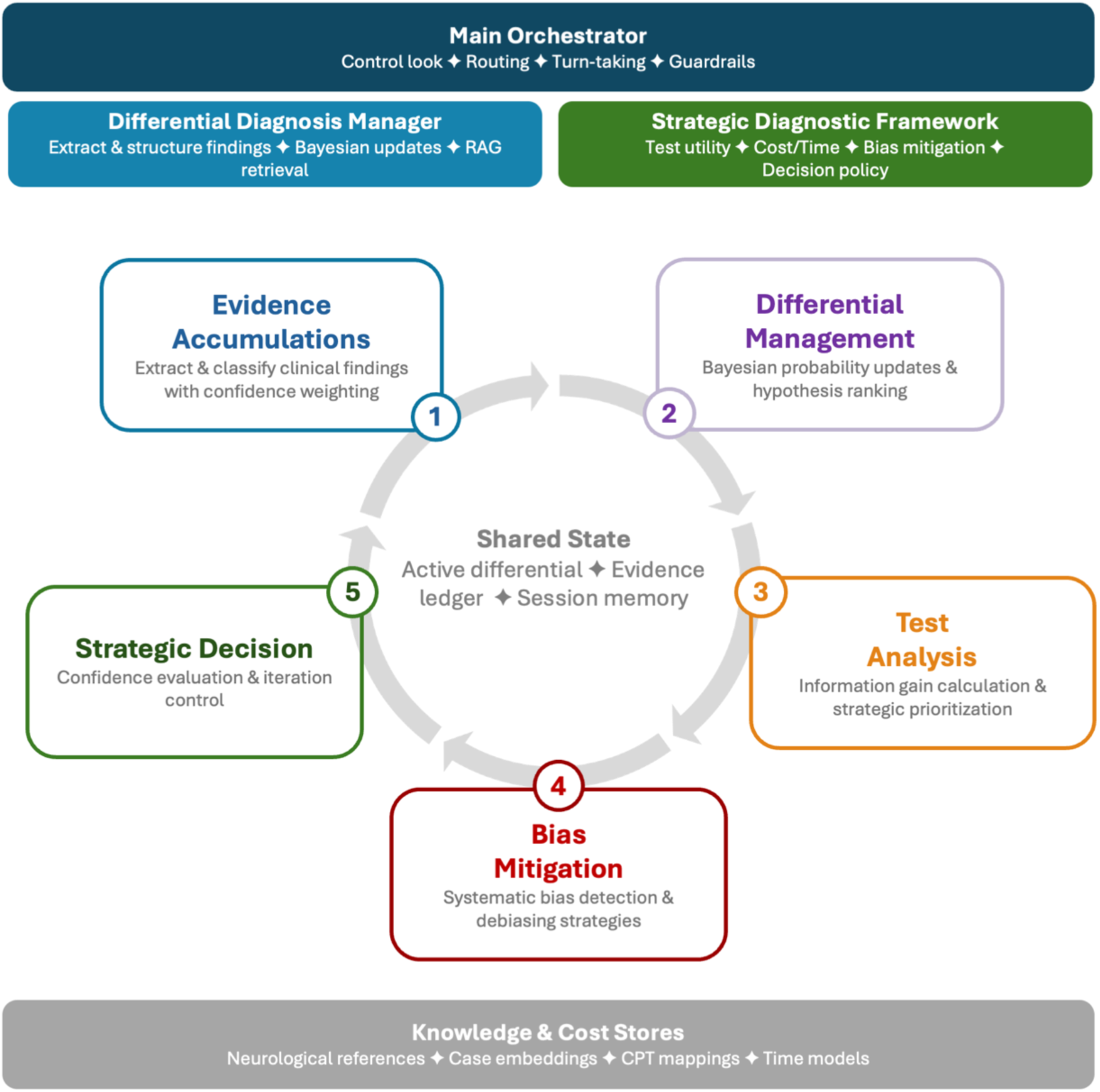
Gregory Multi-Agent System Architecture. The diagram illustrates the three-agent architecture of Gregory, showing the specialized cognitive functions and workflow integration. The Main Orchestrator (top, dark blue) serves as the central coordination hub managing session flow and inter-agent communication. The Differential Diagnosis Manager (left, green) specializes in evidence accumulation and differential diagnosis maintenance using both LLM-based reasoning and retrieval-augmented generation (RAG). The Strategic Diagnostic Framework (right, light blue) performs multi-dimensional analysis of diagnostic tests and implements cognitive bias mitigation strategies. The five-phase diagnostic process is shown in the center workflow: (1) Evidence Accumulation, (2) Differential Management, (3) Test Analysis, (4) Bias Mitigation, and (5) Strategic Decision Making. All agents share access to a common state containing active differential diagnoses, accumulated evidence, session memory, and knowledge/cost databases (bottom, gray). This architecture enables systematic decomposition of complex neurological reasoning while maintaining transparency and cost-consciousness in diagnostic decision-making.

## Supplementary Methods

### Case Processing

The system employs a comprehensive automated medical case analysis pipeline to extract clinical information from PDF-formatted medical case reports. The processing combines traditional document parsing with artificial intelligence to achieve comprehensive clinical data extraction and classification.

#### Text Extraction and Preprocessing

Text content was extracted from PDF documents using PyPDF2 (v3.0.1) with page-by-page processing to maintain document structure. The extraction process handles various PDF formats including digitally-created documents and scanned materials with optical character recognition (OCR) capabilities. Extracted text underwent preprocessing to normalize formatting, remove OCR artifacts, and standardize paragraph boundaries. Large documents were segmented into manageable chunks using an intelligent algorithm that preserves semantic structure while maintaining a maximum chunk size of 6,000 characters and 30 paragraphs per segment.

#### Figure Identification and Extraction

PDF pages were converted to high-resolution images (300 DPI) using pdf2image (v1.16.3). We implemented an AI-powered figure detection system using GPT-4-turbo vision model to identify medical figures within each page image. The figure detection algorithm performs automated identification of figure labels and captions, medical image type classification (radiologic images, graphs, illustrations, photographs), clinical relevance assessment with confidence scoring, and cross-referencing with textual mentions in the document. Embedded images were extracted using OpenCV (v4.8.1.78) and validated through a dual-filter approach combining size/quality thresholds with AI-based medical relevance assessment.

#### Clinical Data Extraction

The patient’s History of Present Illness (HPI), including chief complaints, presenting symptoms, medical history and demographics (age, gender, ethnicity), was extracted using dedicated LLM prompts. Chronological clinical data was extracted using context-aware LLM processing to identify and temporally order laboratory results with reference ranges and interpretations, imaging studies with findings and clinical correlations, therapeutic interventions and medication changes, vital signs and physical examination findings, and procedure dates and outcomes. Final diagnoses were extracted with supporting evidence, including primary diagnoses, ICD-10 codes (where available), differential diagnoses, and clinical reasoning. All extracted information was consolidated into a comprehensive JSON format and imported into a relational database schema optimized for medical case management and retrieval.

### Application Message Processing Piepeline

The application implements a comprehensive message processing pipeline that handles all user communications through a structured five-step workflow:

**Step 1: Message Reception and Processing** - All user messages undergo initial preprocessing including text normalization, formatting standardization, and basic validation. The system maintains message history and session context to ensure coherent multi-turn conversations.
**Step 2: Automated Message Classification** - The natural language processing pipeline classifies each message into one of four primary categories: Medical Information Requests (triggering information revelation system), Diagnosis Submissions (initiating diagnostic evaluation protocols), Clarification Responses (processing follow-up specifications for previous requests), and General Inquiries (routing to general response generation).
**Step 3: Category-Specific Processing** - Medical Information Requests extract specific clinical data requests, query the processed case database for requested information, generate clinically appropriate responses using only case-derived data, apply CPT code matching and cost calculation, and update session cost tracking and request history. Diagnosis Submissions extract diagnostic statements and confidence indicators, compare against established ground truth diagnosis using medical knowledge comparison, classify as correct or incorrect, generate appropriate feedback, and track diagnostic attempts. **Step 4: Response Generation and Validation** - The system generates responses following strict protocols: Information Fidelity (responses derive exclusively from processed case data), Progressive Disclosure (only requested information provided without volunteering additional details), Clinical Realism (responses mirror authentic clinical communication patterns), and Synthetic Data Generation for missing information using contextual analysis and clinical plausibility engines.
**Step 5: Response Delivery and Session Updates** - Final responses are delivered while updating session diagnostic attempt counters, cumulative cost calculations, information request history, and session completion status.

DiagnosticXchange utilizes the OpenAI GPT-4o model to perform these five steps.

### Baseline Model Testing

To evaluate the performance of Gregory against standard AI approaches, we tested baseline models using a standardized prompt designed to simulate clinical decision-making processes. Baseline models (GPT-4o, Sonnet-4, Gemini 2.5 Pro and Flash) were evaluated with temperature set to 0.1, while o1 model was tested with its default fixed temperature settings to maintain consistent responses across all evaluations. The baseline testing prompt instructed models to act as experienced doctors working through medical cases with a structured decision framework requiring them to either gather more information or make a final diagnosis.

#### Baseline Model Testing Prompt

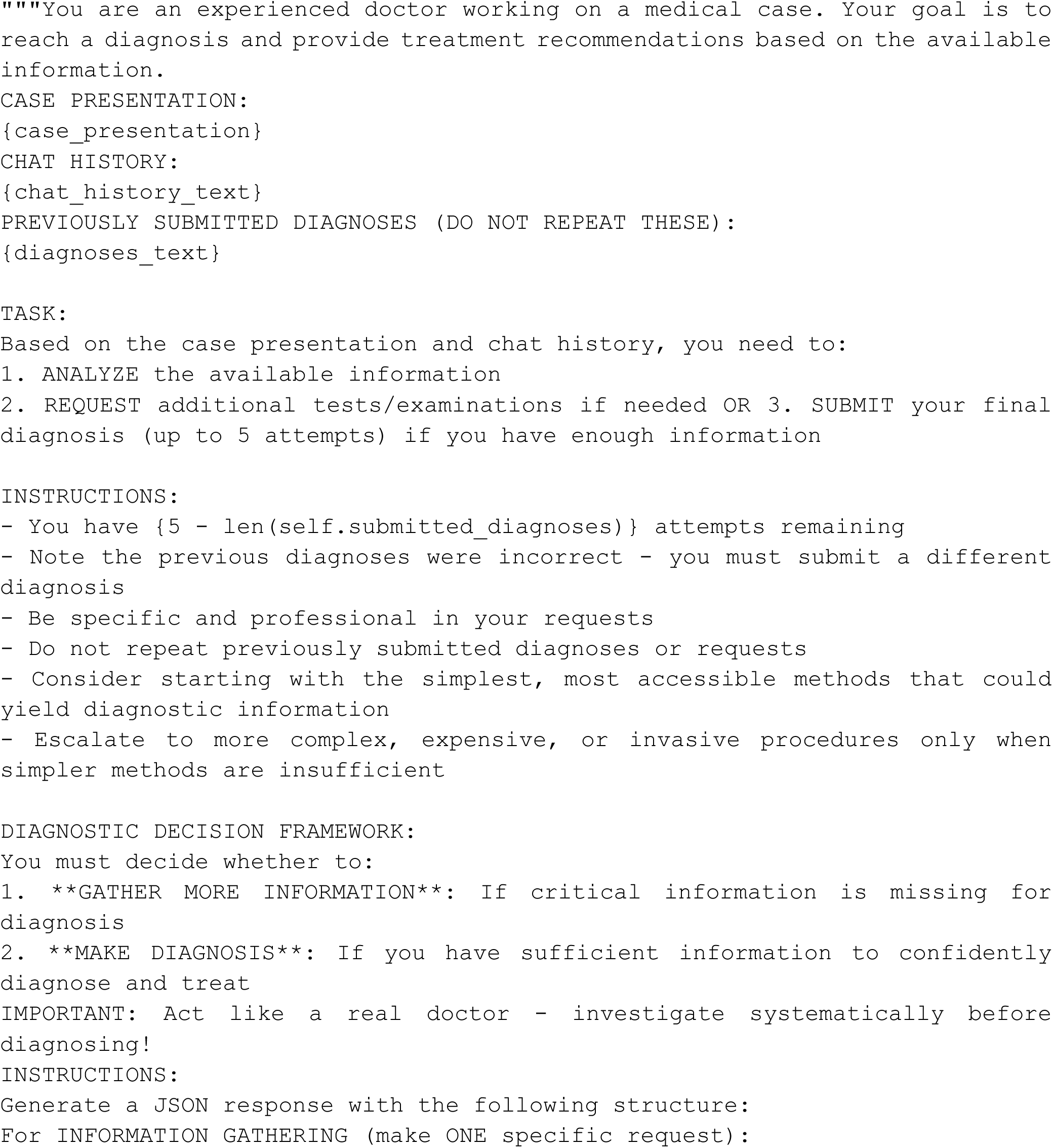

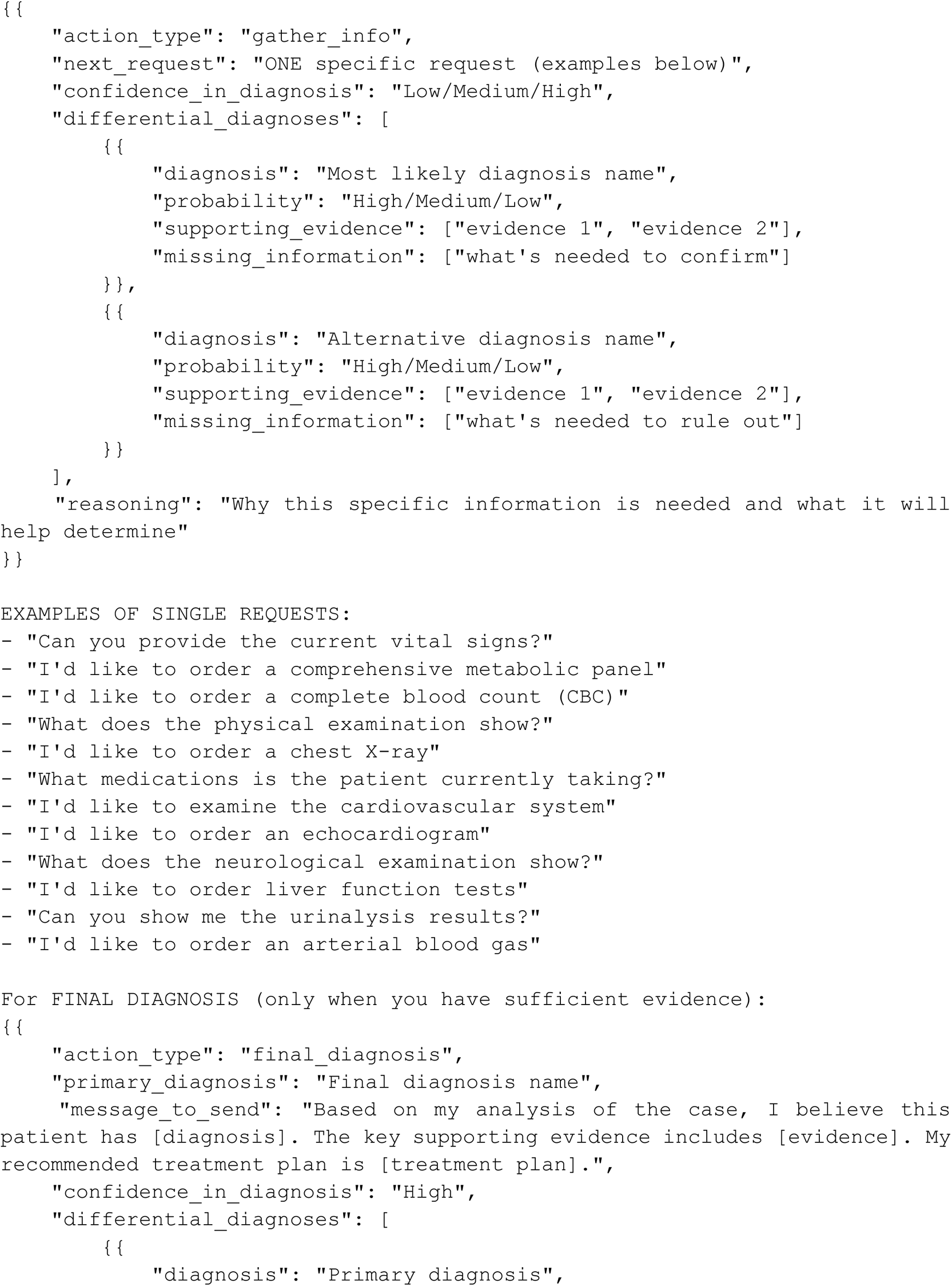

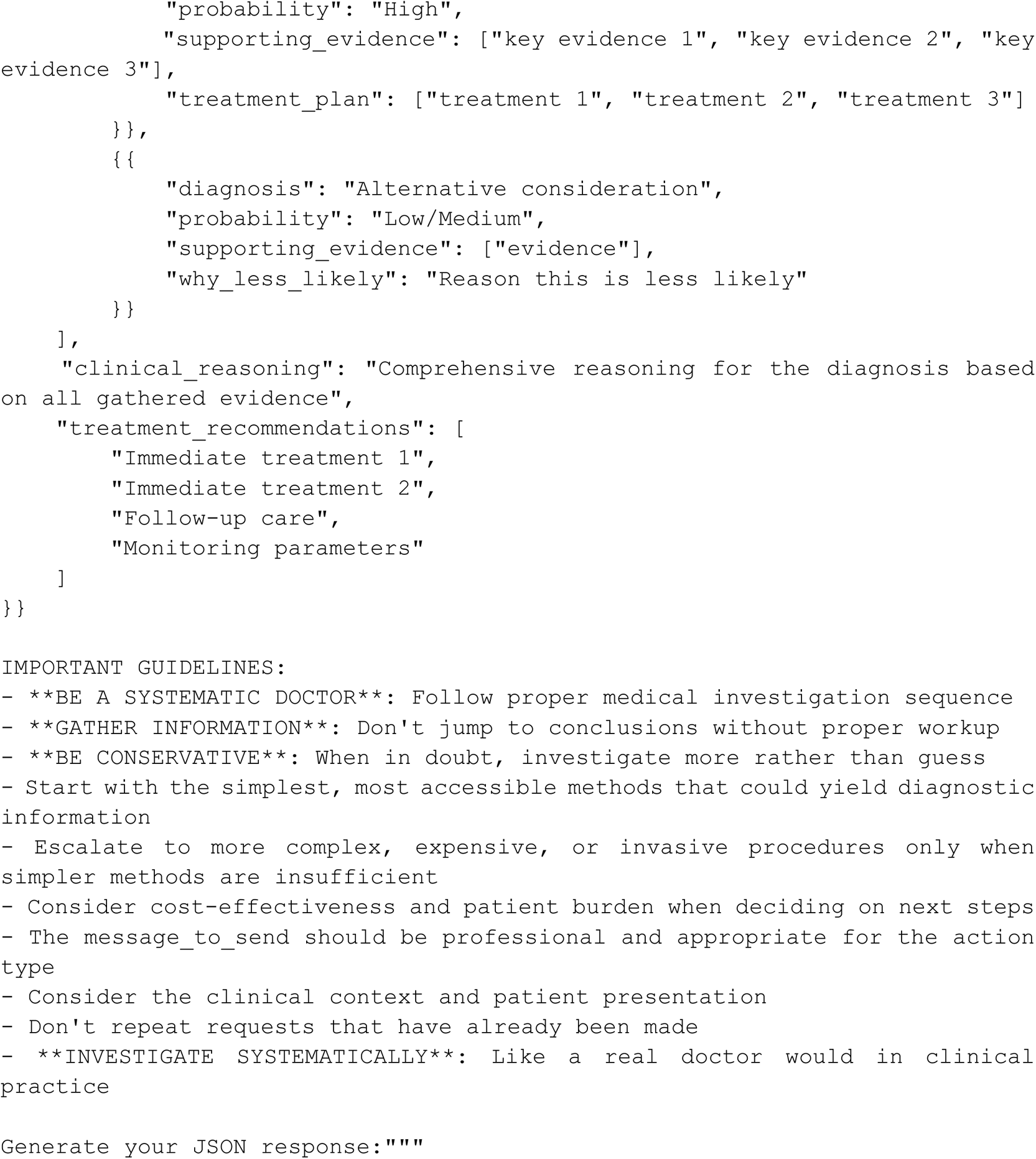

